# The effect of primary health care on AIDS incidence and mortality: a cohort study of 3.4 million Brazilians

**DOI:** 10.1101/2023.10.02.23296417

**Authors:** Priscila FPS Pinto, James Macinko, Andréa F Silva, Iracema Lua, Gabriela Jesus, Laio Magno, Carlos AS Teles Santos, Maria Yury Ichihara, Mauricio L Barreto, Corrina Moucheraud, Luis E Souza, Inês Dourado, Davide Rasella

## Abstract

**Background:** Primary Health Care (PHC) is essential for the health and wellbeing of people living with HIV/AIDS. This study evaluated the effects of one of the largest community-based PHC programs in the world, the Brazilian Family Health Strategy (FHS), on AIDS incidence and mortality.

**Methods:** A retrospective cohort study carried out in Brazil, from January 1 2007 to December 31 2015. We conducted a quasi-experimental effect evaluation using a cohort of 3,435,068 ≥13 years low-income individuals who were members of the 100 Million Brazilians Cohort, linked to AIDS diagnoses and deaths registries. We evaluated the effect of FHS on AIDS incidence and mortality and comparing outcomes between residents of municipalities with no FHS coverage with those in municipalities with full FHS coverage. We used multivariable Poisson regressions adjusted for all relevant municipal and individual-level demographic, socioeconomic, and contextual variables, and weighted with inverse probability of treatment weighting (IPTW). We also estimated FHS effect by sex and age, and performed a wide range of sensitivity and triangulation analyses.

**Findings:** FHS coverage was associated with lower AIDS incidence (rate ratio [RR]:0.76, 95%CI:0.68–0.84) and mortality (RR:0.68,95%CI:0.56–0.82). FHS effect was similar between men and women, but was larger in people aged ≥35 years old both for incidence (RR 0.62, 95%CI:0.53–0.72) and mortality (RR 0.56, 95%CI:0.43– 0.72).

**Conclusions:** AIDS should be an avoidable outcome for most people living with HIV today, and our study shows that FHS coverage could significantly reduce AIDS incidence and mortality among low-income populations in Brazil. Universal access to comprehensive healthcare through community-based PHC programs should be promoted to achieve the Sustainable Development Goals of ending AIDS by 2030.

**Funding:** Funded by the National Institute of Allergy and Infectious Diseases - NIAID/NIH, Grant Number: 1R01AI152938.

## Introduction

In 2021, about 38.4 million people were living with HIV (PLWH) globally, with 650,000 people dying from AIDS-related illnesses.^1^ Strengthening Primary Health Care (PHC) has been identified as a key strategy to achieve the goal of ending AIDS by 2030.^2^ PHC is an integrated health services that, when well-designed, and with universal and equitable coverage, provide comprehensive and coordinated care, including prevention and treatment for people living with HIV/AIDS (PLWHA).^3,4^ HIV prevention takes place in PHC settings through voluntary counseling and testing, health promotion, and the prescribing of antivirals as pre- and post-exposure prophylaxis (PrEP and PEP) for non–HIV-infected patients.^4^ Community-based PHC can promote the use of condoms and safe-sex behaviors, increase HIV testing cost-effectively, enabling early diagnosis, access and adherence to antiretroviral therapy (ART) resulting in reductions in HIV transmission, increasing ART uptake, leading to better viral suppression and retention in care, lower incidence and lower mortality in low- and middle-income countries (LMICs) contexts, reducing HIV/AIDS co-infections, comorbidities, and deaths.^5–11^

Moreover, decentralizing services to a countrywide PHC network should enhance recognition of population needs and improve testing uptake and linkage to care among key populations such as men who have sex with men, female sex workers and injecting drug users who have historically had difficulty accessing services through conventional healthcare providers.^12^

The Family Health Strategy (FHS) is the way Brazil’s national health system provides community-based PHC, and is one of the world’s largest and most evaluated PHC nationwide strategies. FHS characteristics include: universal access with no payment for services; coordinated and comprehensive preventive and curative care delivered by a multidisciplinary team of a physician, a nurse, and community health workers (CHW); geographic empanelment of about 3,500 people residing near the health center’s location; and monthly visits by a CHW who often lives in the area.^13^ The FHS carries out rapid tests for HIV in the community, and updated clinical protocols have encouraged monitoring people with HIV at this level of care.^13^ While the FHS has been associated with improved healthcare access and quality as well as better health outcomes^14^ its impact on HIV/AIDS outcomes has never been studied.^15^ Thus, the aim of this study was to evaluate the effect of a nationwide community-based PHC strategy on AIDS incidence and mortality using a unique cohort of 3.4 million lower-income Brazilians.

## Methods

### Study design, population and ethical issues

We conducted a quasi-experimental study based on longitudinal data from a cohort of 3.4 million low-income individuals aged 13 and older followed up for 9 years. These individuals represent a subgroup of the ‘100 Million Brazilians Cohort’ based on a set of inclusion criteria discussed below. The study protocol was previously published.^16^

This study was approved by the research ethics committee of the Federal University of Bahia, under number 41691315.0.0000.5030 (Report No:3.783.920). The ‘100 Million Brazilians Cohort’ is based on the linkage of administrative data and did not require an additional consent form.

### Data sources

The ‘100 Million Brazilians Cohort’ was previously constructed using the Brazilian national database for social benefits—the Cadastro Único (CadUnico)— which comprises the poorest half of the Brazilian population and is used in determining eligibility for social welfare programs.^17^ This cohort was then linked to the National Disease Notification System (SINAN), a registry of notifiable diseases, and the Mortality Information System (SIM), that registers all deaths and their causes classified by International Classification of Diseases (ICD-10) codes (see Appendix p.2).

These databases were combined using a previously-validated record linkage tool based on the name of the individual and that of their mother, date of birth, sex, and municipality of residence in a two-step procedure.^17,18^ In the first step, the linkage is deterministic. In the second step, entries that were not linked deterministically were then linked probabilistically based on a similarity score between all the pairwise comparisons (i.e., ranging from 0 to 1); entries with the highest similarity scores were considered to be linked pairs.^18^

CadUnico records were linked to municipal FHS coverage data by municipality code with deterministic linkage. Annual FHS coverage was calculated according to the Brazilian Ministry of Health definition: the total number of FHS teams deployed in the municipality in that year multiplied by 3,450—representing the average number of individuals served by each FHS team—divided by the population of the municipality. (Appendix p. 2-3)

The complete Cohort contains information on 114,008,306 individuals. We applied several inclusion criteria to develop our analytical sample as follows: 1) dates - excluding those with registration dates in CadUnico or AIDS diagnosis outside of the study period from Jan 1, 2007 to Dec 31, 2015 (the time period for which national AIDS data were available); 2) treated population - those individuals resident in municipalities where the FHS had 100% coverage over all the study period; 3) comparison (untreated) population - those individuals residing in municipalities where public health facilities provided care via the FHS for less than 20% of the population over all the study period. We used the threshold of <20% coverage for the comparison group - instead of lower coverage levels – in order to: a) capture a higher number of individuals (considering the very low number of municipalities with zero FHS coverage over the all study period), b) given the evidence from the literature that low incipient FHS coverage is poorly effective,^19^ c) because these thresholds has been used by similar individual-based PHC evaluation studies,^20^ and d) because FHS needs a municipal coverage greater than 70% to realize its potential according to previous studies.^19^ However, lower thresholds have been tested in the sensitivity analyses. Moreover, our study population was selected based on 4) Data availability - excluding individuals without complete information for all covariates; and 5) transmission - excluding individuals infected with HIV through vertical transmission.

### Outcomes

Outcomes included: i) AIDS incidence, i.e., new AIDS cases, defined by adapted Centers for Disease Control and Prevention (CDC) criteria, Rio de Janeiro/Caracas criteria, and Death Criterion;^21^ and ii) AIDS mortality rate, i.e., deaths where the underlying cause of death was ICD-10 codes B20 to B24.^21^ Each measure was divided by person-years at risk and multiplied by 100,000. Follow-up time was calculated from the date of entry into the cohort until diagnosis of AIDS, death from AIDS, death from any cause, or the end of the cohort (December 31, 2015), whichever came first.

### Statistical analyses

We performed descriptive analyses of the study population by FHS coverage for each outcome and for selected independent variables. These included demographic characteristics (sex and age), socioeconomic status (skin color, per capita household expenditures, educational attainment, home construction material, number of individuals per room), year of entry into the cohort, time of receipt of the Brazilian conditional cash transfer program Bolsa Família (in months, 0 if the family was not a beneficiary), and annual cumulative AIDS incidence for each individual’s municipality of residence from the study cohort, used as proxy of the municipal-level endemic burden of AIDS.

We selected these variables based on a conceptual framework of the main structural determinants of HIV infection, AIDS incidence and mortality, and of the hypothesized effects of PHC on each step of the evolution of the disease (Appendix, p. 4). Associations between FHS exposure and AIDS incidence and mortality rates were estimated using multivariable Poisson regression models adjusted for all these relevant individual and municipal-level demographic and socioeconomic confounding variables, including follow-up time as an offset variable, with observations weighted using stabilized truncated inverse probability of treatment weighting (IPTW), a consolidated method to evaluate the impact of PHC, and other interventions, with the 100 Million Brazilians Cohort.^20^

We first calculated the IPT weights using multivariable logistic regression models that take into account the probability that each individual lives in a municipality with 100% FHS coverage. Independent variables of the regression included all the baseline demographic and socioeconomic characteristics cited above. Considering Pt as the marginal probability of treatment in the population and PSmul as the propensity score obtained from the multivariable logistic regression adjusted for all independent variables, we calculated the IPTW using the formula (1 – Pt) / (1 – PSmul) for non-treated individuals and used the formula Pt / PSmul for treated individuals. As is standard practice, we dropped extreme weights that were below the first percentile or above the 99th percentile. We then adjusted the IPT weighted multivariable Poisson regression using the same demographic and socioeconomic covariates used to calculate the first stage weights along with: household economic conditions (water supply, type of lighting, garbage disposal and sewage), and municipal conditions (unemployment rate, hospital beds per 1,000 inhabitants). Multivariable logistic regression models for the prediction of the propensity scores, and the odds ratios (OR) for each covariate are presented in Appendix, p. 6.

For each model, multicollinearity was assessed using Variance Inflation Factors (VIF). Subgroup analyses included models by sex and age (under or over the mean age, i.e., 13-34 years and ≥35 years), and additional stratifies were used as complementary analysis (Appendix, p. 7).

### Sensitivity analyses

Sensitivity analyses (Appendix p. 8-16) included first, investigating the effect of defining the comparison group as those living in municipalities with either 0% or with 10% FHS coverage over the study period, to test if the threshold used for the untreated individuals (20%) affected our estimates. Second, we performed a negative binomial regression model comparing results to those found using the IPTW Poisson regression model, to evaluate the robustness of results to other regression specifications. Third, we performed the same multivariable Poisson regressions without IPTW to assess the potential of IPTW for obtaining unbiased FHS estimates. Fourth, we conducted IPTW regressions without the municipal covariates to test the influence of municipal context and resources on our results. This analysis helped to identify whether FHS coverage was truly driving the observed association or whether it was simply collinear with other municipal characteristics. Fifth, we fitted the IPTW Poisson regressions only with a subset of municipalities with high quality of vital information – according to consolidated criteria^22^ - to check if the results are different from the main analysis. Sixth, mortality from external causes (ICD-10 codes V01-Y98) was included as a negative control, since the FHS was not expected to influence them. Seventh, marital status was included in the IPTW regressions as adjusting variable -even with its high number of missing values- to verify if it was a relevant confounder. Eighth, to assess the differences between treated and untreated individuals in relation to sexual preferences and behavior, we estimated the percentage of MSM – variable only available in the records of AIDS cases – in the two groups. We also did complementary tests dividing the analysis in 2 periods, i.e., up to 2010 and after 2011, to check possible differences related to changes in treatments and protocols over the period. We chose for 2011 because in this year PHC started carrying out rapid HIV tests^12^ (Appendix p. 17-24). We also used the records of HIV cases from 2014, year in which they started to be compulsorily notified, to assess if FHS exposure was also associated with potential HIV reductions (even if available study period was of only 2 years). Finally, we performed a triangulation test^23^, using Cox regression (survival analysis) to estimate the FHS effects on AIDS incidence, and compared these findings with our main results (Appendix p. 24). All analyses were performed using STATA version 15.0.

### Role of the funding source

The funding source had no role in study design, data collection, data analysis, data interpretation or the writing of the report.

## Results

We studied 3,435,068 individuals— 605,890 lived in municipalities with zero (or very low) FHS coverage and 2,829,178 lived in municipalities with full FHS coverage during the entire study period (Fig 1). As presented in table 1, although most of the demographic and socioeconomic variables were relatively similar between the two groups, there were also differences in some variables: compared to untreated municipalities, treated municipalities had more men, people of pardo skin color, lower percentage of people with per capita expenses greater than 1 minimum monthly wage, less individuals with more than 9 years of schooling, with garbage disposal and with sewage.

**Figure 1.**
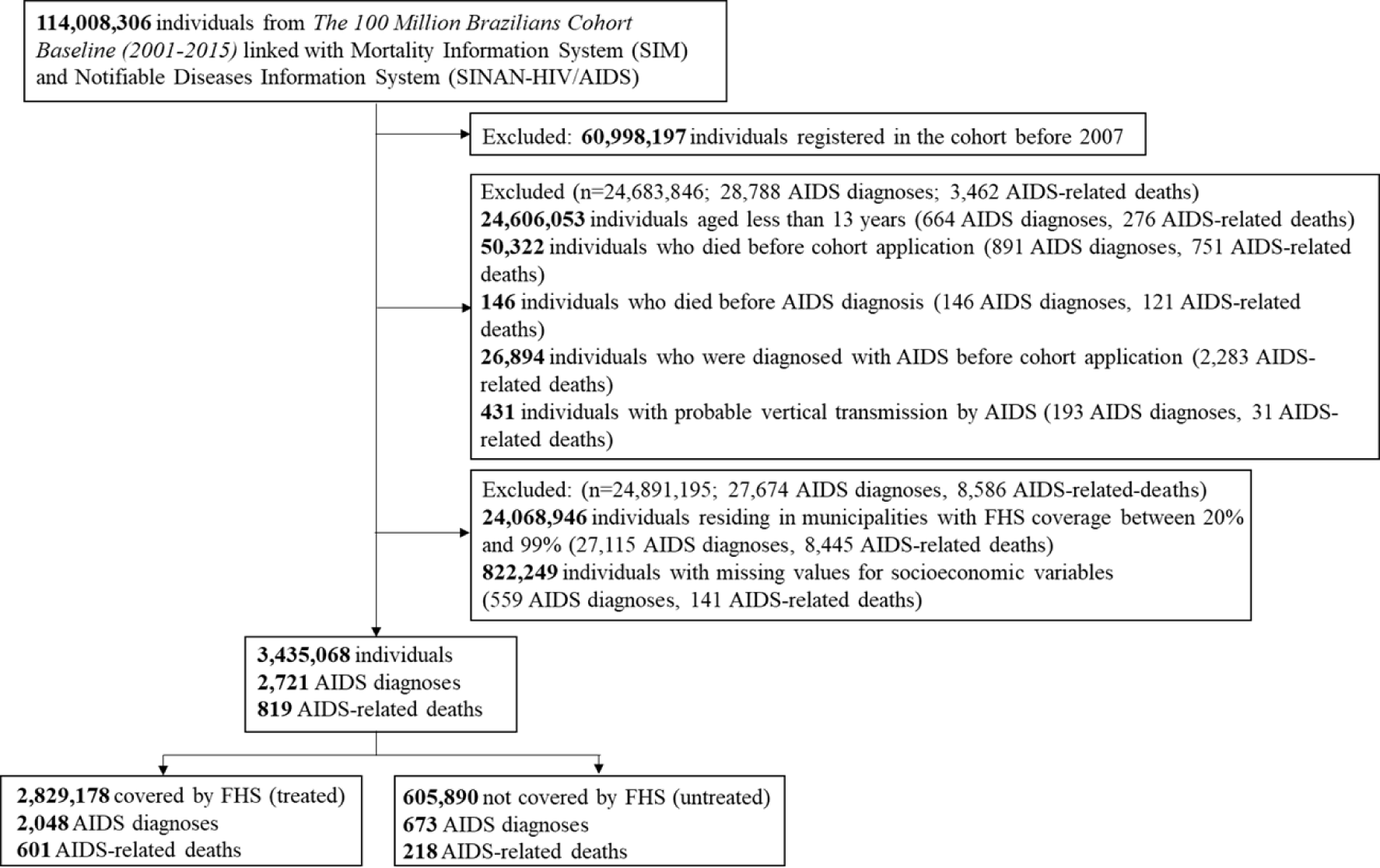
Selection flowchart of the study cohort, The 100 Million Brazilians Cohort, 2007-15

**Table 1.** Individuals treated and untreated by the Family Health Strategy, The 100 Million Brazilians Cohort, Brazil, 2007–15

| Variables | Untreated<br>n=605,890 | Treated<br>n=2,829,178 |
| --- | --- | --- |
| Sex |  |  |
| Female | 335,123 (55.3) | 1,313,027 (46.4) |
| Male | 270,767 (44.7) | 1,516,151 (53.6) |
| Age (years) |  |  |
| 13-24 | 164,746 (27.2) | 936,715 (33.1) |
| 25-59 | 353,953 (58.4) | 1,489,187 (52.6) |
| ≥60 | 87,191 (14.4) | 403,276 (14.2) |
| Skin color |  |  |
| White | 330,124 (54.5) | 811,834 (28.7) |
| Asian | 2,252 (0.4) | 12,737 (0.4) |
| Pardo | 230,255 (38.0) | 1,807,417 (63.9) |
| Black | 42,755 (7.1) | 184,566 (6.5) |
| Indigenous | 484 (0.1) | 13,074 (0.5) |
| Per capita household expenditures - % MW |  |  |
| >1 | 125,660 (20.7) | 206,429 (7.3) |
| 0.5-1 | 172,715 (28.5) | 519,210 (18.3) |
| 0.25-0.49 | 78,011 (12.9) | 402,676 (14.2) |
| 0-0.24 | 119,368 (19.7) | 623,509 (22.0) |
| Nothing declared | 110,136 (18.2) | 1,077,354 (38.1) |
| Education (years of study) |  |  |
| >9 | 194,862 (32.2) | 637,528 (22.5) |
| 4-9 | 165,858 (27.4) | 686,521 (24.3) |
| 1-4 | 197,210 (32.6) | 1,003,227 (35.5) |
| Illiterate, never attended school | 47,960 (7.9) | 502,072 (17.7) |
| Water supply |  |  |
| Public network | 535,461 (88.4) | 1,814,420 (64.1) |
| Well, spring or cistern | 70,429 (11.6) | 1,014,758 (35.9) |
| Home construction material |  |  |
| Brick | 558,976 (92.3) | 2,113,608 (74.7) |
| Wood or taipa<sup>1</sup> | 46,914 (7.7) | 715,570 (25.3) |
| Lighting |  |  |
| Electricity | 554,155 (91.5) | 2,504,416 (88.5) |
| No electricity | 51,735 (8.5) | 324,762 (11.5) |
| Number of individuals per room |  |  |
| 1 | 657,772 (94.1) | 3,306,091 (96.3) |
| 1-2 | 36,202 (5.2) | 109,571 (3.2) |
| >2 | 5,056 (0.7) | 15,703 (0.5) |
| Garbage disposal |  |  |
| Public network | 572,471 (94.5) | 1,794,901 (63.4) |
| Burned, buried or another | 33,419 (5.5) | 1,034,277 (36.6) |
| Sewage |  |  |
| Public network | 472,462 (78.0) | 708,550 (25.0) |
| Septic tank | 34,699 (5.7) | 621,743 (22.0) |
| Rudimentary cesspit/ditch or another | 98,729 (16.3) | 1,498,885 (53.0) |
| Year of entry into the cohort |  |  |
| 2007 | 93,461 (15.4) | 867,836 (30.7) |
| 2008 | 45,980 (7.6) | 284,319 (10.1) |
| 2009 | 42,595 (7.0) | 255,916 (9.0) |
| 2010 | 56,117 (9.3) | 213,462 (7.6) |
| 2011 | 75,914 (12.5) | 228,745 (8.1) |
| 2012 | 90,888 (15.0) | 339,253 (12.0) |
| 2013 | 51,864 (8.6) | 228,104 (8.1) |
| 2014 | 82,339 (13.6) | 252,160 (8.9) |
| 2015 | 66,732 (11.0) | 158,883 (5.6) |
| Time receiving Bolsa Família, months | 30.13 (39.70) | 52.27 (54.14) |
| AIDS municipal incidence among individuals in the cohort<sup>2</sup> | 112.45 (100.23) | 70.09 (76.28) |
| Municipal unemployment rate, % | 7.00 (3.08) | 7.70 (4.17) |
| Hospital beds per 1,000 inhabitants | 1.97 (1.56) | 1.96 (1.99) |

CI=Confidence Interval; %MW - Proportional to the baseline minimum wage (MW).
1- Taipa is a construction method that consists of using clay and wood to build houses
2- Annual cumulative AIDS incidence for each individual's municipality of residence from the study cohort

There were 2,721 new cases of AIDS during the study period, the mean AIDS incidence rate over the period was higher (25.6 per 100,000 person-years) among individuals living in municipalities with no FHS coverage, than among individuals living in municipalities with full FHS coverage (13.2 per 100,000 person-years). During follow-up 819 AIDS deaths occurred, in the untreated group the AIDS mortality rate was higher (8.3 per 100,000 person-years) than among the treated group (3.9 per 100,000 person-years) (Table 2).

**Table 2.** AIDS incidence and mortality rates according to Family Health Strategy (FHS) coverage, Brazil, 2007–15

| Outcome | Untreated<br>N=605,890 | Treated<br>N=2,829,178 | Total<br>N=3,435,068 |
| --- | --- | --- | --- |
| AIDS incidence rate per 100,000 person-years (95%CI) | 25.57 (23.71-27.58) | 13.21 (12.65-13.80) | 15.00 (14.45-15.58) |
| AIDS mortality rate per 100,000 person-years (95%CI) | 8.28 (7.25-9.45) | 3.88 (3.58-4.20) | 4.51 (4.22-4.83) |

CI=Confidence Interval

FHS coverage was associated with a reduction in AIDS incidence (RR 0.76, 95% CI 0.68–0.84), and – with a greater magnitude – with a reduction in AIDS-related mortality (RR 0.68, 95% CI 0.56–0.82) (Table 3). Demographic and socioeconomic adjusting variables showed an overall risk effect of being aged 25-59 years, pardo and black skin color, having a lower level of education, and, having a lower per capita expenditure, while a protective factor for AIDS incidence was being aged ≥60 and reside in a home whose garbage disposal was in the public network (Table 3). The cumulative hazard curves of AIDS incidence and mortality indicates that there was a high and prolonged protective effect of FHS coverage, mainly for AIDS mortality (Appendix, p. 24).

**Table 3.** Inverse probability of treatment weighting Poisson regression models, adjusted for all demographic and socioeconomic variables, for the association between AIDS incidence and mortality rate and Family Health Strategy coverage in the study cohort, Brazil, 2007–15

| Variables | AIDS incidence<br>n=3,435,068<br>RRa (CI 95%) | AIDS mortality<br>n=3,435,068<br>RRa (CI 95%) |
| --- | --- | --- |
| FHS coverage |  |  |
| <20% | 1 | 1 |
| 100% | 0.76 (0.68-0.84) | 0.68 (0.56-0.82) |
| Sex |  |  |
| Female | 1 | 1 |
| Male | 1.11 (1.02-1.21) | 1.15 (0.98-1.34) |
| Age (years) |  |  |
| 13-24 | 1 | 1 |
| 25-59 | 1.97 (1.79-2.17) | 3.52 (2.86-4.32) |
| ≥60 | 0.40 (0.31-0.52) | 0.95 (0.63-1.43) |
| Skin color |  |  |
| White | 1 | 1 |
| Asian | 1.43 (0.74-2.77) | 1.73 (0.55-5.42) |
| Pardo | 1.38 (1.25-1.52) | 1.51 (1.25-1.82) |
| Black | 1.72 (1.49-1.99) | 1.83 (1.42-2.36) |
| Indigenous | 1.08 (0.54-2.17) | 0.51 (0.07-3.63) |
| Education (years of study) |  |  |
| >9 | 1 | 1 |
| 4-9 | 1.52 (1.34-1.72) | 2.41 (1.85-3.14) |
| 1-4 | 1.42 (1.25-1.61) | 2.32 (1.78-3.02) |
| No education | 1.27 (1.08-1.48) | 2.26 (1.65-3.11) |
| Per capita expenditures - % MW |  |  |
| >1 | 1 | 1 |
| 0.5-1 | 1.45 (1.16-1.82) | 1.86 (1.18-2.95) |
| 0.25-0.49 | 1.79 (1.41-2.27) | 1.99 (1.22-3.26) |
| 0-0.24 | 2.00 (1.57-2.56) | 2.67 (1.62-4.39) |
| Nothing declared | 2.21 (1.71-2.83) | 2.79 (1.67-4.63) |
| Home construction material |  |  |
| Brick | 1 | 1 |
| Wood or taipa<sup>1</sup> | 1.18 (1.07-1.31) | 1.10 (0.91-1.33) |
| Water supply |  |  |
| Public network | 1 | 1 |
| Well, spring or cistern | 0.89 (0.80-1.00) | 0.85 (0.69-1.04) |
| Lighting |  |  |
| Electricity | 1 | 1 |
| No electricity | 1.18 (1.04-1.34) | 1.15 (0.91-1.46) |
| Garbage disposal |  |  |
| Public network | 1 | 1 |
| Burned, buried or another | 0.65 (0.57-0.73) | 0.70 (0.56-0.89) |
| Sewage |  |  |
| Public network | 1 | 1 |
| Septic tank | 0.81 (0.71-0.91) | 0.80 (0.64-1.01) |
| Rudimentary cesspit/ditch or another | 0.90 (0.81-1.00) | 0.89 (0.73-1.07) |
| Number of individuals per room |  |  |
| 1 | 1 | 1 |
| 1-2 | 1.02 (0.82-1.26) | 1.28 (0.84-1.95) |
| >2 | 0.78 (0.47-1.31) | 0.73 (0.27-2.01) |
| Year of entry into the cohort |  |  |
| 2007 | 1 | 1 |
| 2008 | 1.14 (1.01-1.29) | 1.01 (0.81-1.26) |
| 2009 | 1.03 (0.90-1.18) | 1.02 (0.80-1.30) |
| 2010 | 1.16 (1.00-1.34) | 0.92 (0.70-1.20) |
| 2011 | 1.12 (0.93-1.36) | 1.02 (0.72-1.45) |
| 2012 | 1.27 (1.05-1.53) | 1.13 (0.81-1.58) |
| 2013 | 1.40 (1.11-1.78) | 1.09 (0.68-1.75) |
| 2014 | 1.45 (1.11-1.88) | 1.07 (0.63-1.81) |
| 2015 | 0.98 (0.59-1.65) | 0.18 (0.02-1.31) |
| AIDS municipal incidence among individuals in the cohort<sup>2</sup> | 1.00 (1.00-1.00) | 1.00 (1.00-1.00) |
| Time receiving Bolsa Família, months | 1.00 (1.00-1.00) | 1.00 (0.99-1.00) |
| Municipal unemployment rate | 1.00 (0.99-1.01) | 1.01 (0.98-1.03) |
| Hospital beds per 1,000 inhabitants | 1.05 (1.03-1.07) | 1.06 (1.03-1.09) |

RRa=rate risk adjusted by sex, age, skin color, education, per capita expenditures, home construction material, number of people per room, year of entry into the cohort, time receiving Bolsa Família (in months), AIDS municipal incidence in the cohort, water supply, lighting, sewage, garbage disposal, municipal unemployment rate, hospital beds per 1,000 inhabitants
1- Taipa is a construction method that consists of using clay and wood to build houses
2- Annual cumulative AIDS incidence for each individual's municipality of residence from the study cohort
CI=Confidence Interval; FHS= Family Health Strategy; %MW - Proportional to the baseline minimum wage

In the subgroup analyses (table 4), the FHS association with AIDS incidence was slightly larger among women (RR 0.70, 95% CI 0.61–0.82), but was significantly stronger for people aged 35 and older (RR 0.62, 95% CI 0.53–0.72). Regarding AIDS mortality, the FHS association was slightly larger among men (RR 0.64, 95ut % CI 0.49–0.83), and significantly stronger among people aged 35 and older (RR 0.56, 95% CI 0.43–0.72) (Table 4).

**Table 4.** Inverse probability of treatment weighting Poisson regression models, adjusted for all demographic and socioeconomic variables, for the association between AIDS incidence and mortality and Family Health Strategy coverage by sex and age, Brazil, 2007–15

| Variables | AIDS Incidence<br>RRa (95%CI) | AIDS Mortality<br>RRa (95%CI) |
| --- | --- | --- |
| Sex |  |  |
| Female | 0.70 (0.61-0.82) | 0.71 (0.54-0.93) |
| Male | 0.79 (0.69-0.91) | 0.64 (0.49-0.83) |
| Age (years) |  |  |
| 13-34 | 0.83 (0.72-0.96) | 0.77 (0.58-1.03) |
| ≥35 | 0.62 (0.53-0.72) | 0.56 (0.43-0.72) |

RRa=rate risk adjusted for sex, age, skin color, education, per capita expenditures, home construction material, number of people per room, year of entry into the cohort, time receiving Bolsa Família (in months), AIDS municipal rate in the cohort, water supply, lighting, sewage, garbage disposal, municipal unemployment rate, and hospital beds per 1,000 inhabitants.
CI=Confidence Interval

All sensitivity analyses confirmed the robustness of the findings, and all triangulation analyses showed a high degree of confidence in the impact estimation (Appendix, p. 8-24).

## Discussion

To the best of our knowledge, this is the first study that evaluates the effect of a nationwide, community-based PHC strategy on AIDS incidence and mortality in a low- and middle-income country. Using a cohort of 3,435,068 individuals and 2,721 new AIDS cases with a robust quasi experiment design, we found a strong effect of FHS coverage on the reduction of both AIDS incidence and mortality. Moreover, FHS showed a stronger impact among individuals ≥35 years old.

These findings add to evidence that the FHS is a robust community-based PHC model associated with improved population health outcomes.^19,20^ Another recent quasi-experiment using a similar cohort and design demonstrated that the FHS was also negatively associated with tuberculosis morbidity and mortality.^20^

Strengthening and expanding PHC are recommended strategies to end AIDS by 2030.^3^ The rationale is that PHC is highly accessible, especially for those with greater social vulnerability,^2^ who are heavily affected by AIDS incidence and mortality.^24^ PHC can foster greater trust with patients and by serving as the entry point to the health system can better facilitate access to HIV diagnosis through rapid testing, linkage to treatment services and enable early initiation of ART, all of which are directly related to the reduction of AIDS incidence.^2,25,26^ HIV prevention actions carried out in PHC, such as sex education, family planning with condom distribution, follow-up and counseling of pregnant and postpartum women living with HIV/AIDS, distribution of syringes and needles to reduce harm among injecting drug users and offering PrEP and PEP, can play an essential role in reducing the incidence of HIV and its progression to AIDS.^3,27^ It is important to point out that PrEP and PEP were introduced in Brazil in 2018, therefore its effects on the treatment were not evaluated in the present study.^28^

PHC should be the ideal place for managing AIDS, since like other chronic conditions it requires maintaining long-term treatments^2^ as well as dealing with other health problems unrelated to HIV, but that come with the increased life expectancy of PLWH. Centralized services that focus only on HIV/AIDS services may increase stigma among people living with the disease^12^ and are limited in their ability to provide comprehensive and integrated care. PHC services, like the FHS, establish long-term ties with users and serve as their medical home, which can increase adherence to AIDS treatment and reduce mortality.^29^ In the FHS, regular home visits, carried out by CHWs, are an important tool for monitoring ART adherence, helping individuals to gain access to other health and social services, identifying other behavioral or environmental risk factors, and delivering tailored education interventions.^29^ A previous study carried out in Brazil showed that the introduction of rapid testing combined with ART–regardless of the patient’s immunosuppression–reduced AIDS incidence and mortality by 60% and 73%, respectively.^28^

The effects of PHC on AIDS incidence and mortality were stronger among older people, a result that was also expected since older people in Brazil tend to have greater access to PHC services.^30^ In Brazil, AIDS mortality is higher among the elderly,^31^ which makes the results of this study even more relevant. However, it is noteworthy that we also found a relevant effect of the FHS in reducing AIDS incidence among people aged 13-34 years. Due to their higher vulnerability to HIV/AIDS, younger populations are a national priority for preventive actions in Brazil.^27^ Our findings are particularly relevant, given that there are studies showing that youths linked to care initiate ART and achieve viral suppression at lower rates than older adults, due to their lower levels of interest in communicable and noncommunicable disease care (which is less prevalent among younger people) and competing priorities, such as those related to social, educational and economic advancement, that outweighed younger adults’ health concerns, resulting in lower care engagement.^32^ On the other hand, when adolescents and youth receive counseling regarding ART adherence, as well as support and referral for psychosocial problems, they had better retention in ART.^33^

### Strengths and limitations of this study

This study has limitations. First, the positive effects of the FHS found in this study are not representative nationally, because all individuals included in the study were derived from the 100 Million Brazilians Cohort, which contains individuals of lower socioeconomic status. Even so, the AIDS incidence rate found in the total population of our study (15.0/100,000 inhabitants, 95%CI:14.4-15.6) was very similar to that of the country in 2021 (16.5/100,000 inhabitants).^21^ Second, we estimated the effect of FHS on AIDS outcomes by adjusting models for multiple socioeconomic variables, nonetheless some unobserved confounders (such as specific characteristics of individuals or variations across municipalities) may not have been accounted for in propensity score-based models. For this reason, we incorporated a wide range of individual-level and municipal-level factors as covariates in regression analyses, and we included the endemic levels of AIDS in the municipality as an adjusting variable that represent non-observed factors associated with the local burden of AIDS before the implementation of the intervention. Results from sensitivity analyses showed that the inclusion of other variables related to inequalities, infrastructure, and healthcare assistance at the municipal level do not affect the FHS effect estimates (Appendix p. 8). Moreover, variables related to sexual behavior, such as marital status and self-identification as Men who have Sex with Men (MSM), which were not included in the main models because of the elevated number of missing values, were nevertheless equally distributed among treated and untreated groups and indicated a predominant heterosexual HIV transmission (self-declared MSM were 13.0% among all AIDS cases) in our cohort of low-income individuals, the percentage of MSM among the linked AIDS cases was particularly low compared to the national proportion of 42.9% in 2021.^21^ Furthermore, when incorporated in the regression models they did not change the direction or magnitude of the results (Appendix p. 11-14). Third, our outcome variables were AIDS-related incidence and mortality (and not HIV incidence) because mandatory notification for HIV only started in 2014 in Brazil.^27^ However, complementary analyses showed a significant negative association between FHS coverage and HIV incidence in 2014-15. Forth, to avoid sparse data, we defined “untreated” individuals as those living in municipalities with <20% FHS coverage. However, sensitivity analyses showed no relevant changes to our results when the untreated groups were defined as those living in municipalities with 0% or <10% FHS coverage (Appendix p. 8). Finally, while the FHS has been found to be a robust model of PHC, not all FHS providers deliver care that is appropriate for some populations at high risk of HIV infection, such as LGBTQ+ individuals. Enhancing the ability of PHC providers to make care more welcoming, appropriate and effective for key populations at risk for HIV infection remains a challenge for the FHS and for PHC in many country contexts.

The main strengths of our study are first its unprecedentedly large cohort of individuals, AIDS cases, and AIDS deaths, and second the robust quasi-experiment design accompanied by a wide range of sensitivity analyses - which confirmed the robustness of the findings - and triangulation tests - that indicate a high degree of confidence in the results.

### Conclusion

In conclusion, our findings show that a universal model of community-based PHC could significantly reduce incidence and mortality from AIDS in LMIC, and even decrease AIDS-related inequities among vulnerable populations. The results of this study justify the expansion and strengthening of effective PHC strategies globally to reach the Sustainable Development Goal of ending AIDS by 2030.

## Declaration of interests

The authors declare no competing interests

## Data Availability

All data supporting the findings presented were obtained from the Centro de Integração de Dados e Conhecimentos para Sauúde (CIDACS). Importantly, restrictions apply to access to the data, which contains sensitive information, were licensed for exclusive use in the current study and, due to privacy regulations from the Brazilian Ethics Committee are not openly available. Upon reasonable request and with express permission from CIDACS (mail to) and approval from an thical committee, controlled access to the data is possible. The dataset is registered under the following DOI handle: https://hdl.handle.net/20. 500.12196/CIDACS/65, which provides metadata and a register of all versions of the database

https://hdl.handle.net/20.500.12196/CIDACS/65

## Acknowledgments

Funding: Funded by the National Institute of Allergy and Infectious Diseases - NIAID/NIH, Grant Number: 1R01AI152938. We acknowledge support from the grant CEX2018-000806-S funded by MCIN/AEI/ 10.13039/501100011033, and support from the Generalitat de Catalunya through the CERCA Program. The funders had no role in the study design or in the decision to submit this manuscript.

We would like to thank all the members of the DSAIDS from Departments of Health Policy and Management and Community Health Sciences, Fielding School of Public Health, University of California (UCLA) for the invaluable discussions and suggestions during the development of this study and the CIDACS members responsible for linkage.

## Data sharing statement

The data underlying this article will be shared on reasonable request to ISC/UFBA and CIDACS Fiocruz and after ethical approval.

## Notes

### Competing Interest Statement

The authors have declared no competing interest.

### Clinical Protocols

https://journals.plos.org/plosone/article?id=10.1371/journal.pone.0265253

### Funding Statement

Funding: Funded by the National Institute of Allergy and Infectious Diseases - National Institutes of Health (NIAID/NIH), Grant Number: 1R01AI152938 awarded to Davide Rasella. We acknowledge support from the grant CEX2018-000806-S funded by MCIN/AEI/ 10.13039/501100011033, and support from the Generalitat de Catalunya through the Centres de Recerca de Catalunya (CERCA Program). The funders had no role in study design, data collection and analysis, decision to publish, or preparation of the manuscript.

